# Burden of Obstetric Fistulas in Zambia: A Bayesian Modelling Approach

**DOI:** 10.64898/2026.08.24.26361281

**Authors:** Mercy M. Imakando, Isaac Fwemba, Kwasi Torpey, Timothy J. Robinson, Emefa Modey, Goshon Kasanda, Charles Michelo, Eugene Kaunda, Ernest Maya, Anthony Danso-Appiah

## Abstract

**Background:** Obstetric fistula, a severe but preventable complication of childbirth, disproportionately affects women in low-resource settings, but its true burden in Zambia remains uncertain owing to inconsistent routine health data collection and limited surveillance. This study aimed to estimate the national and subnational burden of obstetric fistula in Zambia using Bayesian modelling.

**Methods:** The records of women who underwent obstetric fistula repair from nine national Fistula Repair Facilities in Zambia were reviewed from 2018 to 2023 to obtain district-specific and country wide data. District-level birth data was obtained from the national Health Management Information System (HMIS). A Bayesian hierarchical model was implemented to estimate fistula burden, accounting for variation in the number of births and uncertainty in observed case counts. Posterior burden estimates were reported per 1,000 births with 95% credible intervals (CrIs).

**Results:** A total of 928 fistula cases were reported across 4,964,598 births between 2018 and 2023 with an estimated national burden of 0.28 per 1,000 births (95% CrI: 0.25–0.35). Provincial burden ranged from 0.07 per 1,000 births (95% CrI: 0.04–0.13) in Southern Province to 0.61 per 1,000 births (95% CrI: 0.46–0.95) in Northern Province. The highest district-level burden was observed in Chama at 2.48 per 1,000 births (95% CrI: 1.24–4.89).

**Conclusion:** The estimated national burden of obstetric fistula in Zambia is lower than reported from many high burden countries in sub-Saharan Africa, however, considerable district and provincial disparities persist. Strengthening maternal health services and expanding fistula prevention and treatment programs in high-burden areas should be prioritized.

## Introduction

An average of 800 women die daily as a result of pregnancy and childbirth related complications [1]. For each maternal death, roughly 100 other women suffer severe pregnancy-related complications that can result in lifelong disability [2]. This translates to about 30 million women with severe maternal morbidity per annum [3]. Sub-Saharan Africa bears a disproportionate share of this burden [4] with approximately one third of pregnant women suffering from maternal morbidities [5]. Despite the scale of the problem, the true global burden of maternal morbidity remains uncertain due to gaps in measurement and reporting [6]. Understanding the full extent of these morbidities is crucial in informing health system responses and policy interventions. Severe maternal morbidities include uterine prolapse, stress incontinence, hypertension, severe anaemia and fistula [3]. Among these, the obstetric fistula is considered the worst [7].

An obstetric fistula is an abnormal connection between two body cavities or a body cavity and the skin [8]. For obstetric fistulas, the connection is between the vagina and the urinary tract and/or the rectum, occurring as a complication of childbirth [9, 10]. An estimated 100,00 new cases of obstetric fistula develop each year, most of them in Asia and Sub-Saharan Africa (SSA) [11]. The global prevalence of obstetric fistulas is estimated at 0.29 per 1000 women of reproductive age, increasing to 1.60 per 1000 women of reproductive age for women in SSA [12]. In neighbouring Malawi, the prevalence is estimated at 1.6 per 1000 women [13]. In Zambia, the precise extent of the obstetric fistula problem has not been fully characterized. Based on analysis of the 2013-14 Demographic Health Survey (DHS), the prevalence of obstetric fistula in Zambia was estimated at 5.91 per 1000 Women of Reproductive Age [14]. However, diagnosis of obstetric fistula from the DHS was based on the symptoms of obstetric fistula without clinical validation, which like many other studies where the diagnosis is based upon symptoms, is prone to overestimated prevalence [13, 15]. The lack of reliable data on Zambia’s fistula burden represents a critical knowledge gap, one that has implications for health planning and resource allocation.

Accurate estimates of the burden of obstetric fistulas are essential for tracking progress towards elimination, informing the delivery of prevention and treatment services, and guiding surgical training and resource allocation. This study seeks to generate robust national and district level estimates of the burden of obstetric fistula in Zambia. To achieve this, we apply statistical modelling techniques that integrate multiple data sources including health facility case records, the national Health Management Information System (HMIS), and population data from the most recent census [16]. This approach addresses limitations of individual data sources and enables the generation of more accurate and geographically detailed estimates. The findings will provide evidence to inform policies, programs and interventions pertaining to equitable distribution of emergency obstetric and fistula repair service. Furthermore, the study will establish a stronger evidence base for monitoring progress towards the elimination of obstetric fistula and improving health, wellbeing and dignity of women and girls in Zambia.

## Methods

### Study design

The study reviewed medical records to identify women who underwent obstetric fistula repair between 2018 and 2023. These data were combined with district-level birth data obtained from the national Health Management Information System (HMIS) to estimate the burden of obstetric fistula at district and national level. Data were accessed for research purposes between 01/01/2023 and 30/09/2023.

### Study Sites

Primary data was collected from nine health facilities from eight provinces across Zambia that offer fistula repair surgeries. The participating facilities were; Chilenje 1^st^ level Hospital (Lusaka Province), Chilonga Mission Hospital (Muchinga Province), Kabwe General Hospital (Central Province), Mansa General Hospital (Luapula Province), Mbala General Hospital (Northern Province), Monze Mission Hospital (Southern Province), Women and Newborn Hospital (Lusaka Province), Solwezi General Hospital (North-Western Province) and St. Francis Mission Hospital (Eastern Province). Data collection took place from January to September 2023.

The geographic locations of these facilities are shown in Fig 1.

**Fig 1.**
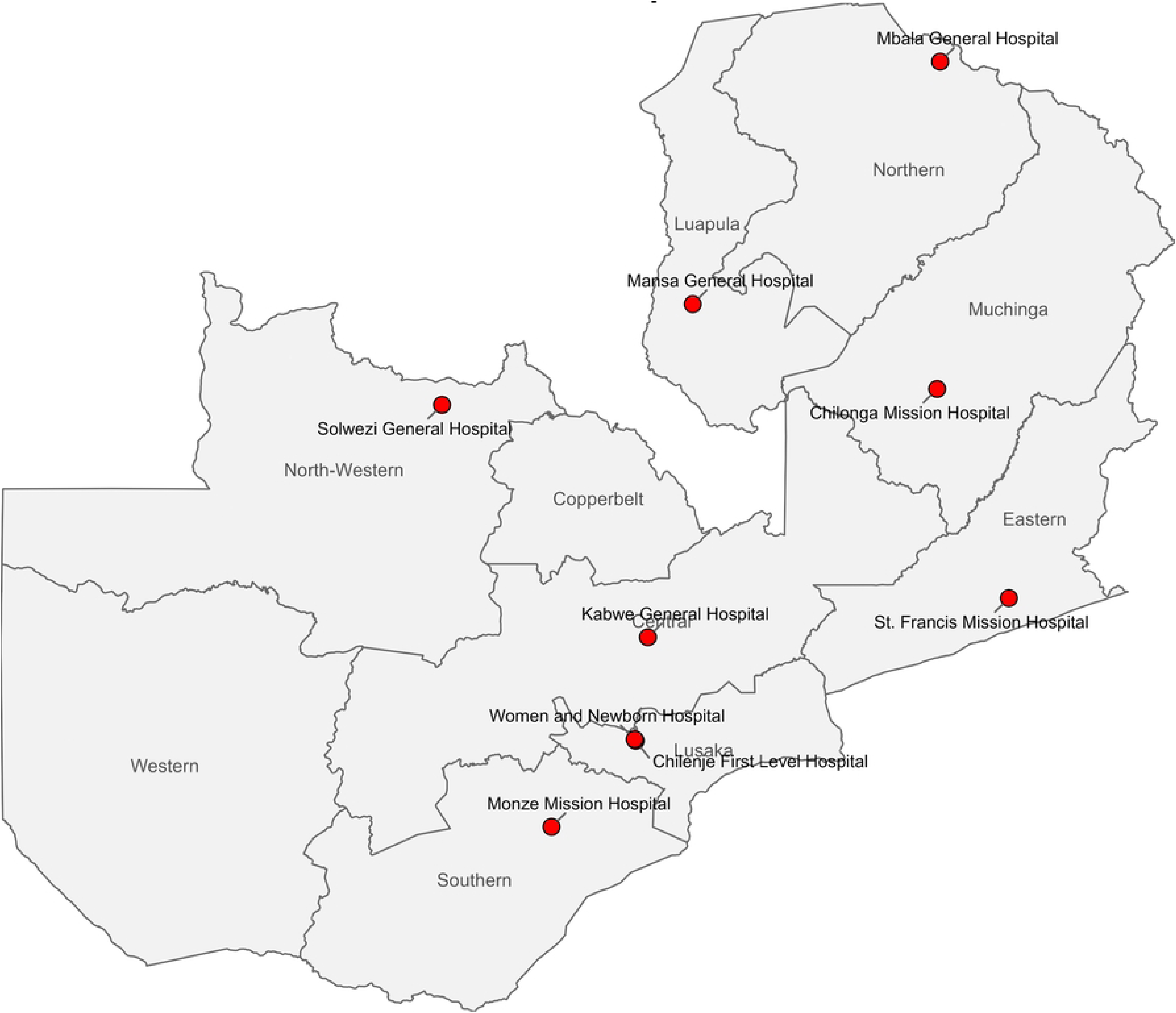
Map of Zambia showing the study sites.

### Patients

Women who underwent obstetric fistula repair between January 2018 and September 2023 were included. Records of women who underwent obstetric fistula repair with missing information on residence were excluded.

### Procedures

For each case identified, data on the women’s age, residence (by district and province), marital status, education levels and past obstetric history were captured and entered onto a predesigned data entry sheet. Complementary secondary data was obtained from the national Health Management and Information System (HMIS) and the 2022 Census of Population and Housing [16]. From the HMIS, information on the number obstetric fistulas and births recorded in each district for the years 2018 to 2023 was extracted. From the 2022 census, demographic information, including the distribution of women by age and population by district and province. These secondary data provided denominators and population context necessary for the calculation of obstetric fistula burden.

To validate the accuracy of obstetric fistula case data reported in Zambia’s Health Management Information System (HMIS), a verbal, Delphi-style consultation was conducted with maternal health experts and data officers at district and provincial levels. Participants included heads of obstetrics and gynaecology, district medical officers, and health information officers. Through informal open-ended interviews, which were conducted in person or via phone, the plausibility of reported obstetric fistula cases from 2018 to 2023 were conducted, to provide contextual explanations for anomalies in the data.

### Statistical Analysis

Statistical analysis was conducted in R. Descriptive statistics were used to summarise the sociodemographic and obstetric characteristics, with categorical variables presented as frequencies and percentages using available data for each variable as the denominator. Delays in obstetric care were assessed at three points: at home before seeking facility care, at the clinic/RHC/health post before referral, and at the hospital before delivery. A delay was defined as ≥ 24 hours at each point.

A Bayesian hierarchical model was applied in estimating district-level obstetric fistula burden. The model accounted for differences in the number of births across districts and allowed information to be shared across districts, thereby stabilising estimates in districts with small populations or few observed cases. The model was implemented in Just Another Gibbs Sampler (JAGS) through the rjags interface in R, allowing full Bayesian inference via Markov Chain Monte Carlo (MCMC) simulation. We modelled the observed number of obstetric fistula cases in each district as a Poisson random variable, with its mean proportional to the number of births in that district.

For district j, let yⱼ denote the observed number of obstetric fistula cases and Bⱼ the total number of births during the study period. The observed case count was modelled as:

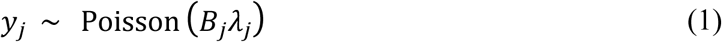

where λⱼ denotes the latent district-level obstetric fistula burden per birth and Bⱼ is the exposure team.

To allow burden to vary by district while still pooling information across all districts, we specified a hierarchical prior for the district-level rates. In the second stage of the model, we assumed that the log-burden in each district follows a common normal distribution:

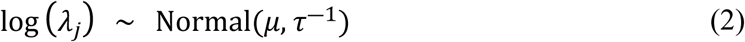

*μ* is the overall (national) mean log-burden and *τ* is the precision (inverse of the variance) of the log-burden. This hierarchical specification results in partial pooling, whereby estimates from districts with sparse data are drawn towards the overall mean to a greater extent than estimates from districts with more information.

Weakly informative priors were chosen for the model’s hyperparameters μ and τ, reflecting minimal prior knowledge so that the data largely drive the inference.

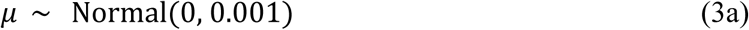

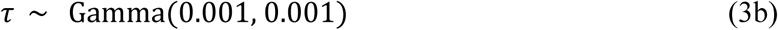

Where the second parameter of the normal distribution denoted precision under the JAGS parameterisation.

Posterior Inference and MCMC Sampling

The two-level Bayesian model was fitted using MCMC sampling in JAGS via rjags in R. Three parallel MCMC chains were run, each for 5,000 iterations after a burn-in of 1,000 iterations, with an adaptation phase of 1,000 iterations. No thinning was applied. Convergence was evaluated using trace plots and the Gelman–Rubin diagnostic (R^), ensuring R^ < 1.1 for all parameters.

For each MCMC iteration s, district leel burden per 1,000 births was calculated directly from the posterior draw of the latent district rate:

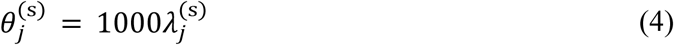

For a province or the nation, represented by a group G of districts, the aggregate burden was calculated as a birth-weighted posterior rate:

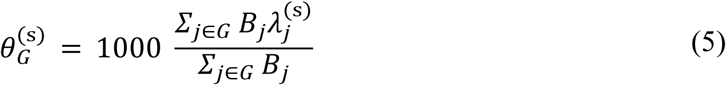

Posterior medians were reported as the point estimates, and 95% credible intervals defined by the 2.5th and 97.5th percentiles of the corresponding posterior distributions.

Posterior predictive counts were generated as:

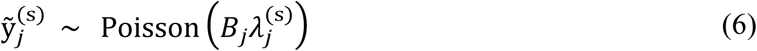

And were used for posterior predictive assessment of model fit rather than as primary basis for estimating the district burden.

All analyses were conducted in R (version 4.5.1). The rjags package was used for model fitting, coda for posterior diagnostics, and dplyr, tidyr, and ggplot2 for data processing and visualization. Spatial mapping of burden estimates was done using the sf and tmap packages.

### Ethical considerations

Ethical approval was obtained from the University of Zambia Biomedical Research Ethics Committee (UNZABREC) REF NO. 3388-2022. Permissions to conduct the research were obtained from the National Research Authority (NHRA) Ref No. NHRA0000015/20/12/2022, Ministry of Health and the heads of participating health facilities. The study involved a retrospective review of anonymized medical records, with direct patient contact or collection of identifiable personal information. Individual informed consent was not obtained because only anonymised routinely collected clinical records were reviewed.

## Results

From January 1, 2018 to September 30, 2023, there were 958 Obstetric fistula surgeries conducted at the nine fistula repair facilities in Zambia. Thirty cases were excluded from the analysis: 24 involved women who were residing outside Zambia (19 from Mozambique, four from Malawi and one from the Democratic Republic of Congo), while six were excluded because of missing data. The present analysis is based on data from the remaining 928 women.

The HMIS data recorded 1318 fistulas. Inconsistencies were noted during the Delphi process. Not all recorded fistulas were clinically validated. In areas without an obstetrician, suspected cases were entered as cases, and the data did not differentiate obstetric from non-obstetric fistulas. As a result of the above, HMIS data was not included in the analysis. Fig 2 below outlines the data collection and screening process.

**Fig 2.**
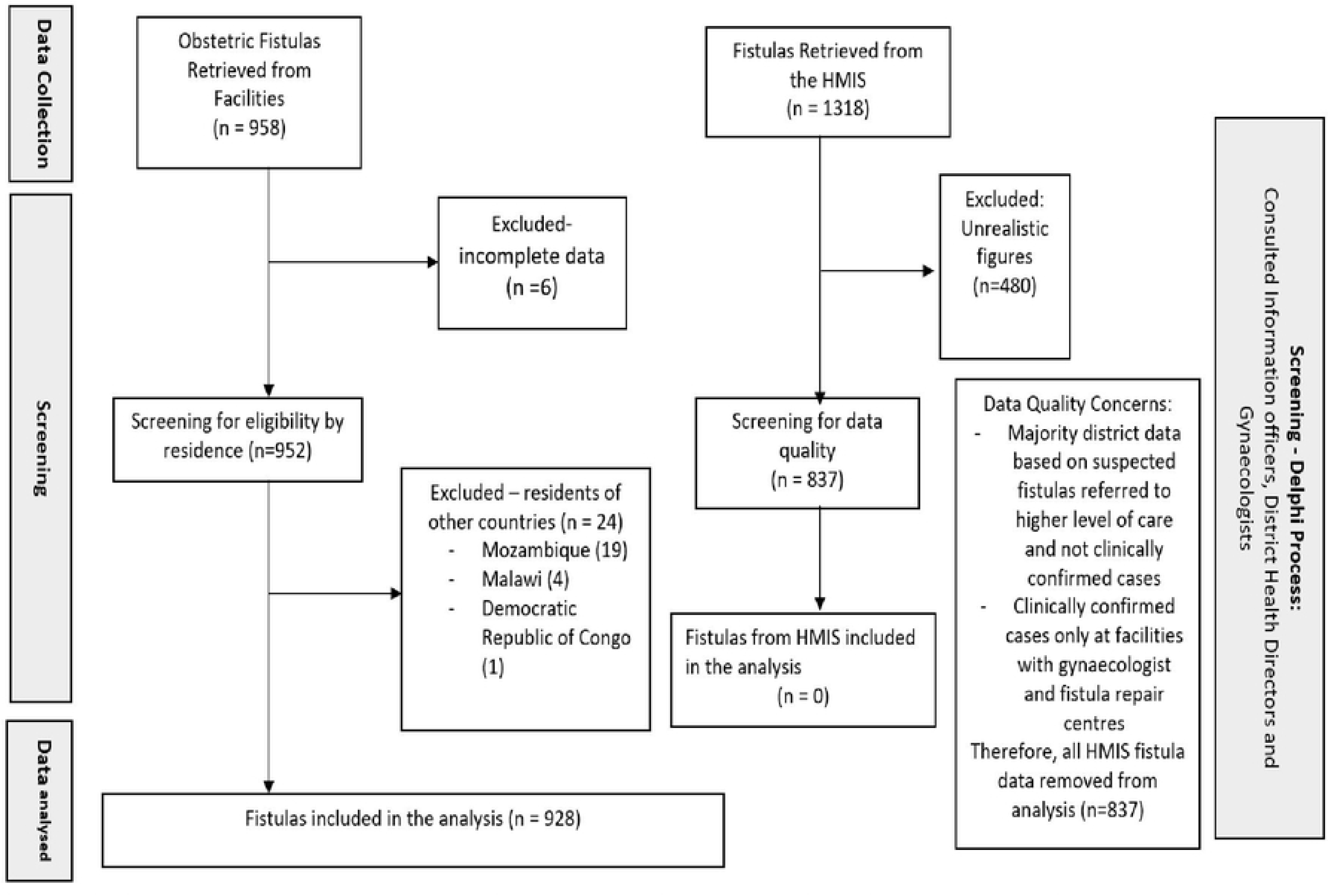
Flow Diagram Indicating Data Collection and Screening of Records for Obstetric Fistulas in Zambia.

### Socio-demographic characteristics of the patients

The median age of the participants at the time of fistula repair was 32 years (IQR 24 – 40), The youngest patient underwent repair at age 17, while the oldest was aged 78. Education levels among the women were low. Just over a quarter (25.9%) had no formal education, while 53.4% had incomplete primary education (Table 1).

**Table 1.** Socio-demographic Characteristics of women with obstetric fistulas in Zamia.

| **Characteristic** | **Frequency**<br>**(n = 928)** | **Percentage** |
| --- | --- | --- |
| **Residence (Province)** |  |  |
| Central | 117 | 12.6 |
| Copperbelt | 29 | 3.1 |
| Eastern | 256 | 27.6 |
| Luapula | 97 | 10.5 |
| Lusaka | 41 | 4.4 |
| Muchinga | 83 | 8.9 |
| North Western | 65 | 7.0 |
| Northern | 194 | 20.9 |
| Southern | 30 | 3.2 |
| Western | 16 | 1.7 |
| **Current Age** |  |  |
| <20 | 82 | 8.8 |
| 20 - 29 | 303 | 32.7 |
| 30 - 39 | 282 | 30.4 |
| 40 - 49 | 151 | 16.3 |
| 50 and above | 97 | 10.4 |
| Not indicated | 13 | 1.4 |
| **Marital Status** |  |  |
| Single | 91 | 9.8 |
| Married | 598 | 64.4 |
| Separated/divorced | 162 | 17.5 |
| Widow | 43 | 4.6 |
| Not Indicated | 34 | 3.7 |
| **Education (n=378)** |  |  |
| No Formal Education | 98 | 25.9 |
| Incomplete Primary | 202 | 53.4 |
| Completed Primary | 14 | 3.7 |
| Incomplete Secondary | 55 | 14.6 |
| Completed Secondary | 4 | 1.1 |
| Completed Tertiary | 5 | 1.3 |
| **Literacy (n=162)** |  |  |
| Not able to read and write | 126 | 77.8 |
| Able to read but not write | 2 | 1.2 |
| Able to write | 34 | 21.0 |

### Obstetric characteristics of participants

Over 2/3^rd^ (68.3%) were multiparous at the time of fistula repair. Most (93.7%) of the participants delivered from health facilities and 56.8% delivered operatively (Table 2). Majority (73.3%) of women with obstetric fistula had a still birth in the pregnancy preceding fistula development.

**Table 2.**
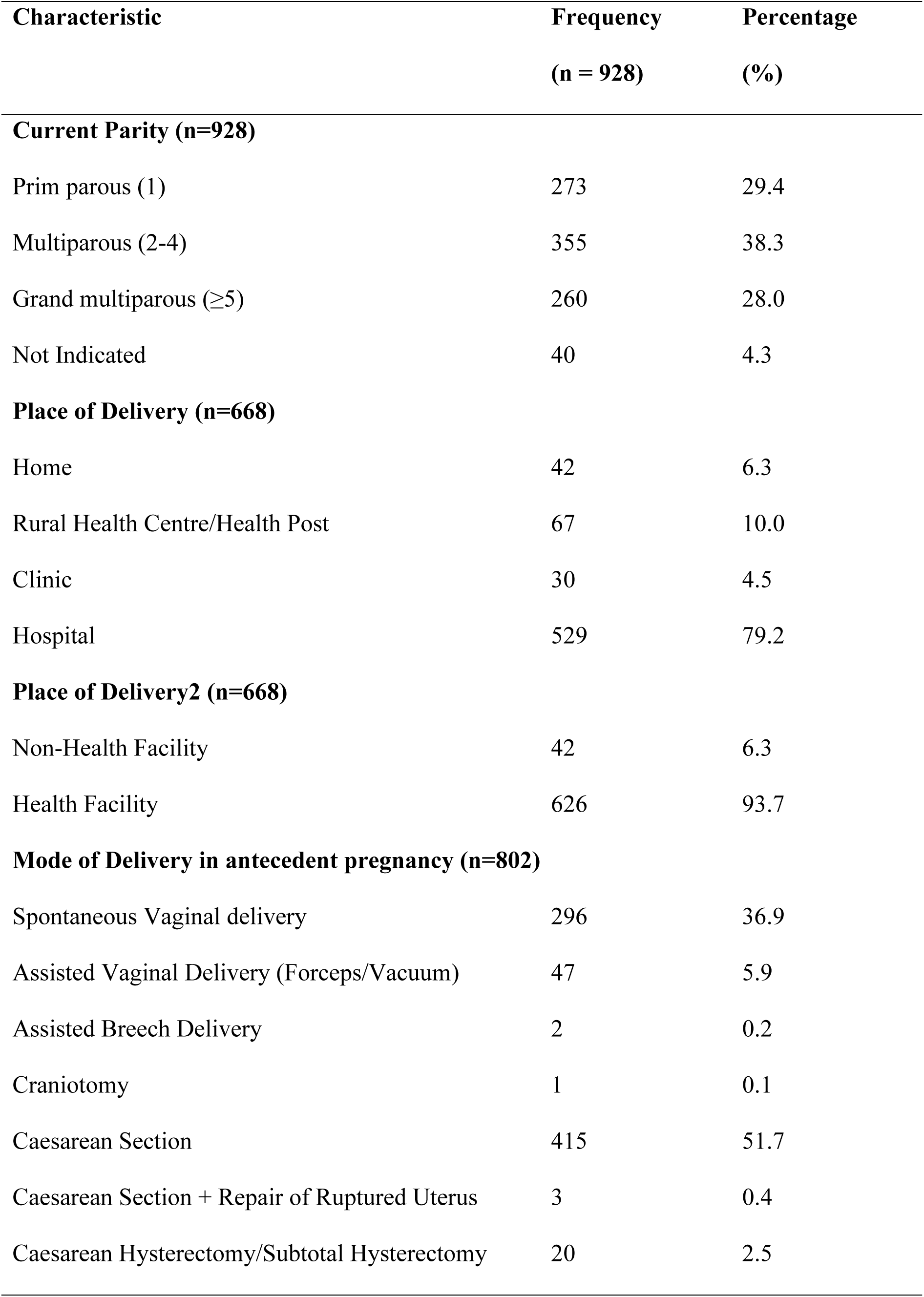
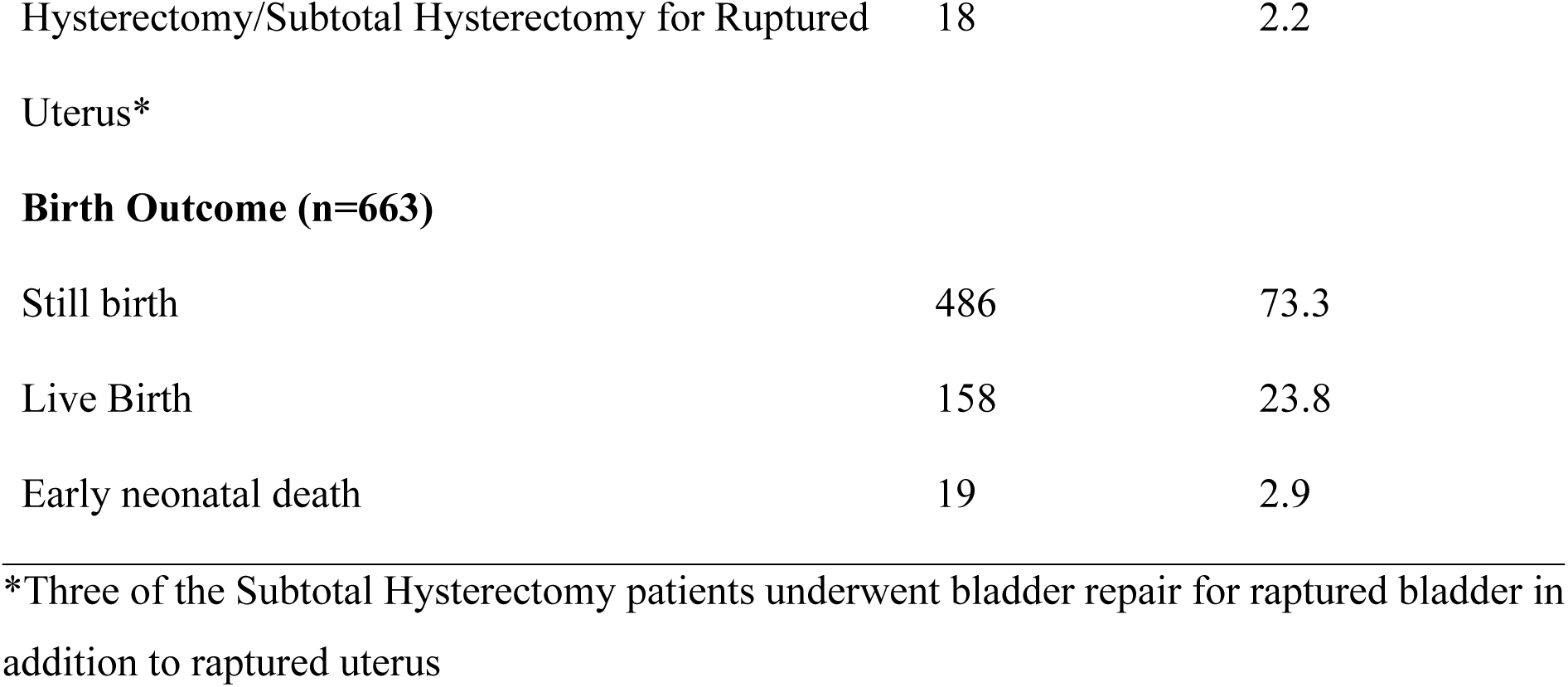
Obstetric characteristics of women with obstetric fistulas in Zambia.

| **Characteristic** | **Frequency**<br>**(n = 928)** | **Percentage**<br>**(%)** |
| --- | --- | --- |
| **Current Parity (n=928)** |  |  |
| Prim parous (1) | 273 | 29.4 |
| Multiparous (2-4) | 355 | 38.3 |
| Grand multiparous ($\geq 5$) | 260 | 28.0 |
| Not Indicated | 40 | 4.3 |
| **Place of Delivery (n=668)** |  |  |
| Home | 42 | 6.3 |
| Rural Health Centre/Health Post | 67 | 10.0 |
| Clinic | 30 | 4.5 |
| Hospital | 529 | 79.2 |
| **Place of Delivery<sup>2</sup> (n=668)** |  |  |
| Non-Health Facility | 42 | 6.3 |
| Health Facility | 626 | 93.7 |
| **Mode of Delivery in antecedent pregnancy (n=802)** |  |  |
| Spontaneous Vaginal delivery | 296 | 36.9 |
| Assisted Vaginal Delivery (Forceps/Vacuum) | 47 | 5.9 |
| Assisted Breech Delivery | 2 | 0.2 |
| Craniotomy | 1 | 0.1 |
| Caesarean Section | 415 | 51.7 |
| Caesarean Section + Repair of Ruptured Uterus | 3 | 0.4 |
| Caesarean Hysterectomy/Subtotal Hysterectomy | 20 | 2.5 |
| Hysterectomy/Subtotal Hysterectomy for Ruptured Uterus* | 18 | 2.2 |
| **Birth Outcome (n=663)** |  |  |
| Still birth | 486 | 73.3 |
| Live Birth | 158 | 23.8 |
| Early neonatal death | 19 | 2.9 |

\*Three of the Subtotal Hysterectomy patients underwent bladder repair for ruptured bladder in addition to ruptured uterus

Most women (93.7%) delivered at health facilities (Table 2). Figure 3 illustrates delays of ≥24 hours at different points in the care pathway according to place of delivery. Among women who delivered at a RHC/health post, 44% experienced a delay at home and 52% at the RHC/health post. Among those who delivered at a clinic, 33% experienced a delay at home and 33% at the clinic/RHC/health post level. Among women who delivered at a hospital, 31% experienced a delay at home, 40% at the clinic/RHC/health-post level before referral, and 29% at the hospital before delivery. These findings demonstrate that substantial delays occurred at multiple points before delivery, particularly at the primary health-care and referral levels. Fig 3).

**Fig 3.**
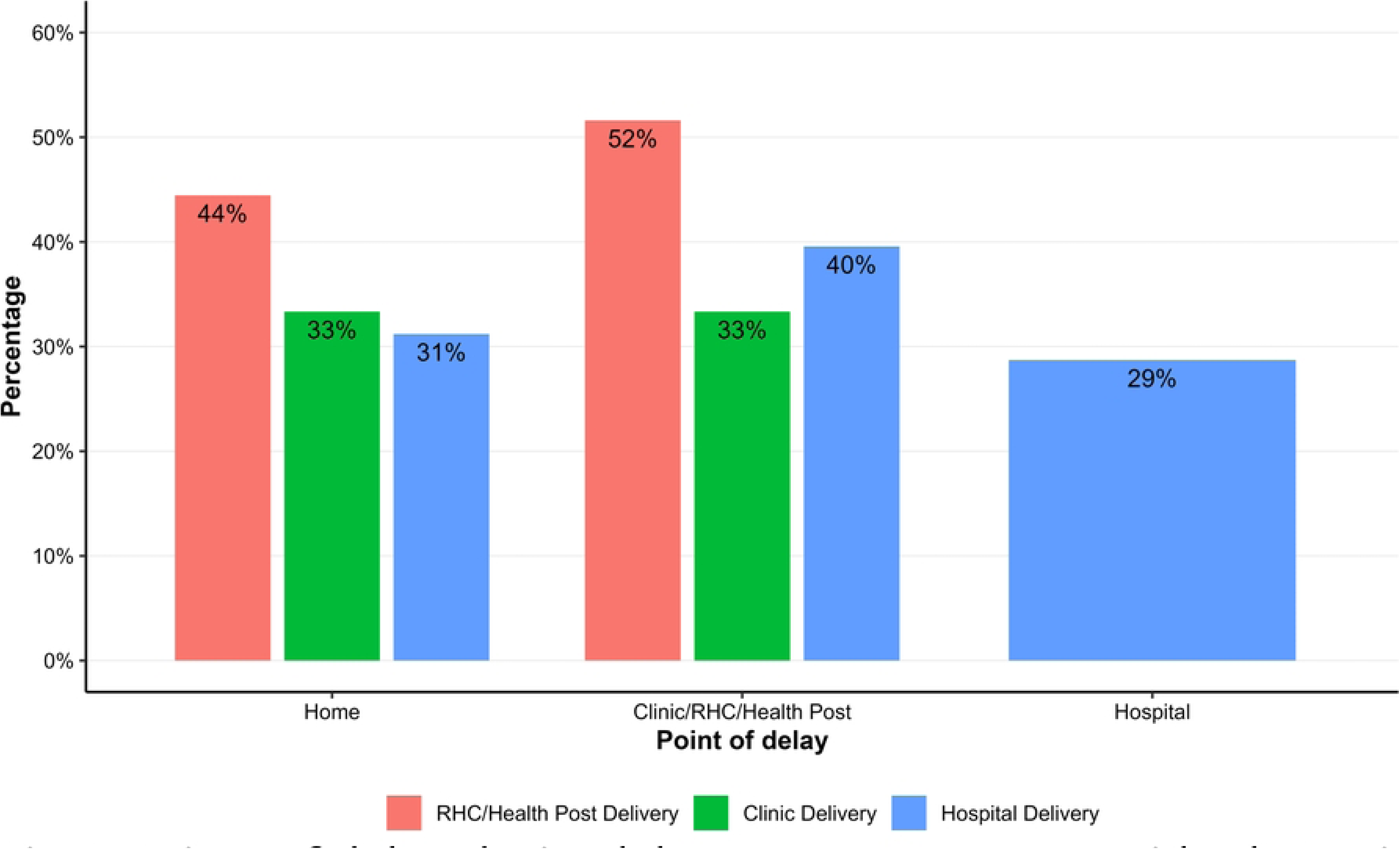
Points of delay during labour among women with obstetric fistula in Zambia 2018 – 2023. Note: Percentages were not cumulative as it was possible for a woman to delay at one or more points of care St Francis Mission Hospital the highest number of obstetric fistulae with 184 women while Women and Newborn hospital recorded the lowest with 14 women (Table 3).

**Table 3.** Number of obstetric fistula by facility and year in Zambia (2018 – 2023)

| Facility | 2018 | 2019 | 2020 | 2021 | 2022 | 2023 | Total |
| --- | --- | --- | --- | --- | --- | --- | --- |
| Chilenje General Hospital | 0 | 27 | 24 | 1 | 17 | 5 | 74 |
| Chilonga Mission Hospital | 55 | 55 | 13 | 12 | 30 | 7 | 172 |
| Kabwe Central Hospital | 17 | 25 | 10 | 22 | 27 | 11 | 112 |
| Mansa General Hospital | 40 | 49 | 15 | 1 | 17 | 16 | 138 |
| Mbala General Hospital | 14 | 39 | 36 | 27 | 0 | 22 | 138 |
| Monze Mission Hospital | 2 | 4 | 6 | 8 | 9 | 4 | 33 |
| Solwezi General Hospital | 0 | 14 | 16 | 0 | 18 | 15 | 63 |
| St Francis Mission Hospital | 0 | 8 | 57 | 32 | 40 | 47 | 184 |
| Women and Newborn Hospital | 7 | 5 | 2 | 0 | 0 | 0 | 14 |
| Total | 135 | 226 | 179 | 103 | 158 | 127 | 928 |

### Burden of obstetric fistulas in Zambia

The estimated national burden of obstetric fistula in Zambia is 0.28 per 1,000 births (95% CrI: 0.25-0.35), approximately 1 obstetric fistula per 3600 births. Variations in the burden are noted among the provinces, ranging from 0.07-0.61 per 1,000 births. The provinces with the lowest burden of obstetric fistula in Zambia were Southern-0.07 per 1,000 births (95% Crl: 0.04–0.12) and Copperbelt-0.07 per 1,000 births (95% Crl: 0.04–0.13). Northern Province had the highest burden of obstetric fistulas at 0.61 per 1,000 births (95% Crl: 0.46–0.89) followed by Eastern Province with 0.53 per 1,000births (95% Crl: 0.41–0.81) (Table 4).

**Table 4.** Burden of obstetric fistula in Zambia (2018 – 2023) by province.

| Province | Obstetric Fistulas | Births (HMIS <sup>a</sup> ) | Burden per 1,000<br>births (95% CrI)<sup>β</sup> |
| --- | --- | --- | --- |
| Northern | 194 | 334,919 | 0.61 (0.46–0.89) |
| Eastern | 256 | 512,162 | 0.53 (0.41–0.81) |
| Muchinga | 83 | 180,623 | 0.48 (0.33–0.76) |
| Central | 117 | 341,392 | 0.35 (0.24–0.55) |
| North Western | 65 | 208,329 | 0.33 (0.22–0.53) |
| Luapula | 97 | 342,885 | 0.28 (0.19–0.44) |
| Lusaka | 41 | 527,938 | 0.08 (0.04–0.16) |
| Western | 16 | 215,988 | 0.08 (0.03–0.15) |
| Copperbelt | 29 | 429,453 | 0.07 (0.04–0.13) |
| Southern | 30 | 428,313 | 0.07 (0.04–0.12) |
| Zambia | 928 | 4,964,598 | 0.28 (0.25–0.38) |

<sup>a</sup>HMIS – Health Management and Information System
<sup>β</sup> Bayesian Posterior Estimates with their 95% credible interval

At district level, the estimated obstetric fistula burden ranged from 0 to 2.48 per 1,000 live births. Districts with the highest burden of obstetric fistula in Zambia were, Chama (2.48 per 1,000 births 95% Crl: 1.24–4.89) in the Eastern Province, Serenje (1.41 per 1,000 births 95%: 0.63–2.89) in Central Province, Nsama (1.38 per 1,000 births 95% Crl 0.49–3.17) in Northern Province, Mpulungu (1.32 per 1,000 births 95% Crl: 0.60–2.57) in Northern Province and Kaputa (1.23 per 1,000 births 95% Crl: 0.44–2.57) in Northern Province (Figure 5.2). Of the 116 districts in Zambia, 15 districts recorded no fistulas. Of these, 53% (n = 8) were from Western Province, while 38% (n = 5) were from Southern Province (Fig 4).

**Fig 4.**
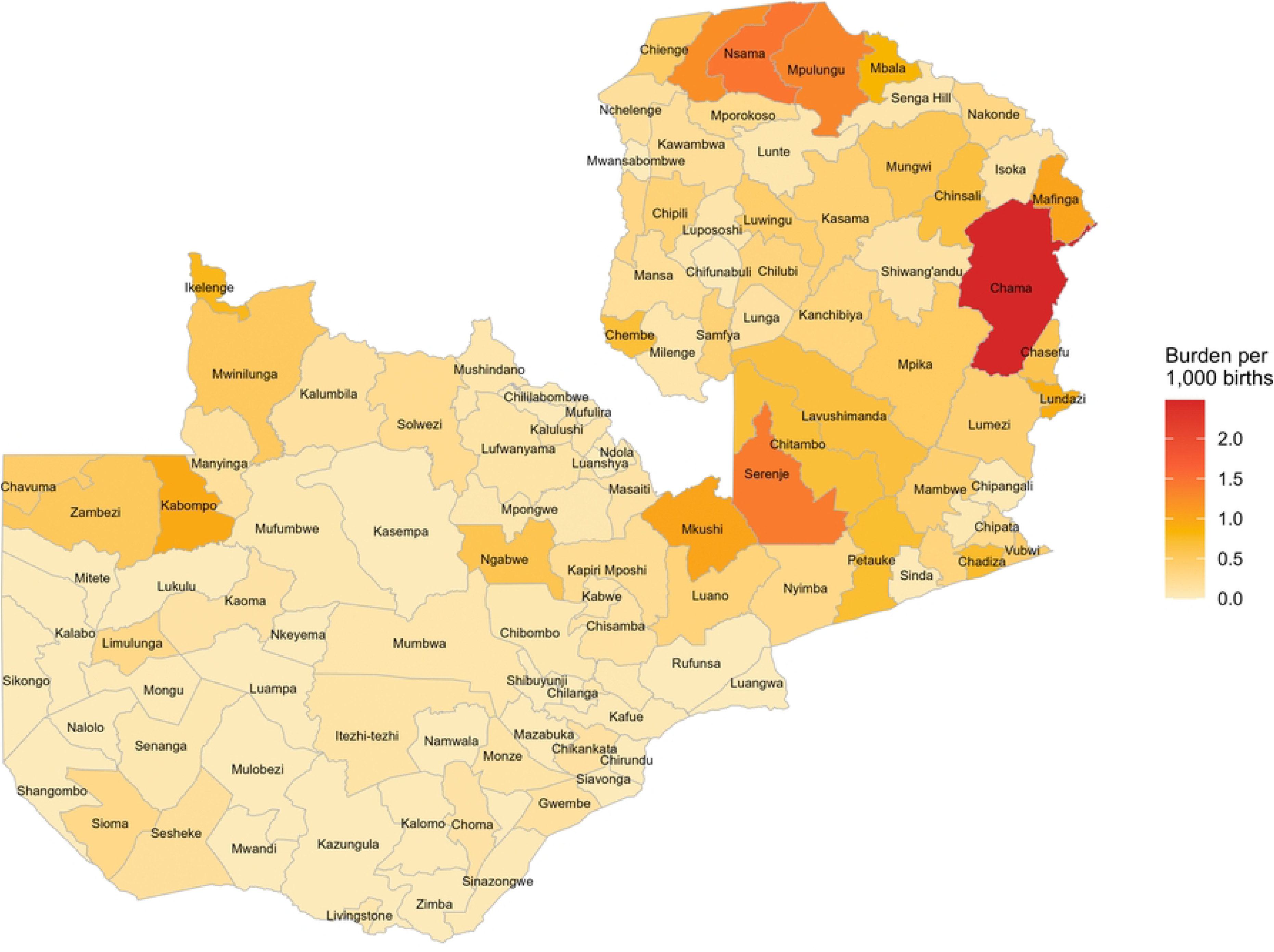
District-Level Burden of Obstetric Fistula per 1,000 Births in Zambia, 2018–2023.

## Discussion

These findings indicate that Zambia’s estimated national burden of 0.28 per 1,000 births, equivalent to roughly one fistula per 3,600 deliveries—falls within the lower range of burden estimates documented in other settings. Global studies, for instance, have documented incidence rates varying from nearly zero in some populations to approximately 4 per 1,000 deliveries in the most affected areas [17]. Community-based studies have reported prevalence estimates ranging from 0.26 to 1.06 per 1,000 women of reproductive age, while facility-based studies have observed rates of approximately 0.51 per 1,000 women of reproductive age [12]. The present findings are also lower than estimates from Ghana (1.6 – 1.8 per 1,000 deliveries) [18], indicating possible contextual and systemic differences in maternal health services, surveillance systems, and case detection efficiency.

These cross-study comparisons must, however, be interpreted with caution due to differences in denominators and study designs. Some prevalence estimates are expressed per 1,000 births or deliveries, whereas others use 1,000 women of reproductive age or 1,000 recently pregnant women as the reference population. Each denominator represents a distinct at-risk group—reflecting different exposure periods and probabilities of obstetric complications—thus complicating direct comparisons. Standardization of measurement frameworks and reporting metrics is therefore essential to improve the comparability and interpretation of obstetric fistula burden data across settings. Despite the low burden, each fistula case represents a childbirth gone devastatingly wrong and carries profound personal and public health significance. Additionally, obstetric fistula at any level is cause for concern and underscores continuing gaps in maternal healthcare.

The distribution of obstetric fistulas across Zambia’s provinces demonstrates pronounced spatial and socioeconomic inequities in healthcare access, consistent with patterns in other resource-limited settings [19, 20]. The burden varied nearly nine fold across provinces, from 0.07 per 1,000 births in Southern and Copperbelt Provinces to 0.61 per 1,000 births in Northern Province. The highest-burden provinces—Northern, Eastern, Muchinga, and Central—are predominantly rural, with rural population proportions exceeding 75% [16]. Conversely, Lusaka and Copperbelt, Zambia’s most urbanized provinces (over 80% urban), recorded the lowest burden, mirroring the protective influence of urbanization and healthcare access.

This rural–urban gradient underscores structural inequities in health system. Limited access to emergency obstetric and newborn care (EmONC) is a key driver. Nationally, only 18.4% of the recommended EmONC facilities are operational [21], reflecting major service delivery gaps. Northern Province, the most affected region, has the lowest facility coverage (7.5%), followed by Muchinga at 13.3%. Although Eastern Province exhibits slightly better EmONC availability (22.9%), its high burden suggests additional contributing factors such as delayed care-seeking, cultural barriers, and early childbearing. These disparities point to systemic failures in the distribution and functionality of obstetric services. Additional disparities were noted in education levels.

Education-related disparities further compound these challenges. Most women affected by obstetric fistula had limited formal education, with 83% attaining only primary level or less. This finding aligns with prior studies [22, 23], which consistently demonstrate that low educational attainment is linked with increased fistula risk. Limited education not only constrains women’s health literacy and awareness of maternal danger signs but also reduces their likelihood of accessing skilled birth attendance and timely emergency care [24].

The district-level findings further emphasize the compounding effects of geography and infrastructure. Chama District in Eastern Province had the highest burden (2.48 per 1,000 births), district is plagued by challenges of poor road infrastructure and low health worker staffing levels, which limit both women’s ability to reach health facilities promptly and their access to skilled care upon arrival [25], amplifying the risk of fistula formation. Similarly, remote Northern districts such as Kaputa, Mpulungu, and Nsama recorded high burden, correlating with documented access challenges. For instance, in Kaputa, the average travel time to a health facility was 105 minutes, and 70% of women travelled on foot [26]. In Western Province, fewer than 20% of women live within two hours of an EmONC facility [27]. These findings exemplify two of the “three delays”, delays in reaching a facility, and receiving adequate treatment—all of which heighten fistula risk [28]. Globally, access inequities are stark: the met need for EmOC is 99% in high-income countries but only 21–32% in low-resource contexts [29].

Obstetric fistula remains both a symptom and a sentinel indicator of inadequate maternal healthcare. Its persistence highlights the need to strengthen Zambia’s health system—particularly in ensuring universal access to quality emergency obstetric care. The findings demand renewed investment in infrastructure, human resources, and referral systems to achieve equitable care distribution. Expanding the number and functionality of EmONC facilities in high-burden areas should be a top priority. Strengthening maternal health workforce capacity, especially in midwifery and surgical obstetrics, will further reduce the delay between labor complications and life-saving interventions.

Zambia’s National Obstetric Fistula Strategic Plan (2022–2026) [30] provides a vital policy framework. The current findings should guide resource allocation, prioritizing Northern, Eastern, Muchinga, and Central Provinces for preventive and curative interventions. Addressing rural transport and infrastructure barriers is equally critical to achieving safe and timely delivery care.

## Conclusion

The estimated national burden of obstetric fistula in Zambia is lower than that reported from high-burden countries in sub-Saharan Africa. However, substantial provincial and district-level disparities persist, reflecting inequities in access to timely, high-quality maternal health care. Eliminating obstetric fistula is achievable but requires sustained investment in emergency obstetric care, fistula prevention and treatment services, and targeted interventions in high-burden areas. Despite the relatively low national burden, the persistence of even a few cases signify that preventable maternal injuries continue to occur, underscoring the need to ensure every woman has access to safe childbirth and the dignity of a life free from obstetric fistula.

## Data Availability

The minimal data set and accompanying code is available at figshare via https://doi.org/10.6084/m9.figshare.33274497

## Acknowledgements

The authors are grateful to the Ministry of Health of Zambia, for granting permission to conduct this study and the participating fistula repair facilities, provincial and district health offices, and the Health Management Information System team for their support and provision of data. We also acknowledge the health records personnel, research assistants, and the women whose anonymized clinical records made this research possible.

